# Insulin-dependence as a Predictor of Shortened Cancer-specific Survival in Pancreatic Neuroendocrine Tumors: A Multi-Institutional Study from the United States Neuroendocrine Study Group

**DOI:** 10.1101/2024.09.11.24313436

**Authors:** Muhammad Bilal Mirza, Jordan J Baechle, Paula Marincola Smith, Danish Ali, Mary Dillhoff, George Poultsides, Flavio G. Rocha, Clifford S. Cho, Emily R. Winslow, Ryan C. Fields, Shishir K. Maithel, Kamran Idrees

## Abstract

**Introduction:** PNETs are rare pancreatic malignancies originating from islet cells and exhibit a strong co-occurrence with Diabetes Mellitus (DM), associated with worse survival outcomes. However, studies have yet to delineate the impact of insulin dependent (IDDM) and non -insulin dependent (NIDDM) on poor oncological outcomes.

**Methods:** Utilizing the U.S. Neuroendocrine Tumor Study Group database (1999-2016), we performed a retrospective cohort study of adult patients who underwent primary surgical resection of PNETs. Patients were categorized based on preoperative diagnosis into non-DM, NIDDM, and IDDM cohorts. We used the Kaplan-Meier method and log-rank test to study cancer-specific survival (CSS). Cox proportional Hazards models were used to assess the impact of IDDM on CSS.

**Results:** Of the 1,122 patients included in the analysis, 870 (77%) were non-DM, 168 (15%) were NIDDM, and 84 (8%) were IDDM. The groups were similar in tumor stage and grade. However, they differed in sex, BMI, age, ASA class, tumor location, preoperative HbA1c and serum glucose (*p*-value <0.05). Patients with IDDM had significantly decreased 5-year CSS compared to patients without IDDM (CSS: IDDM 85%, NIDDM 94%, non-DM 93%, NIDDM + non-DM 93%; *P* <0.01). On multivariate analysis, IDDM was independently associated with worse CSS (HR 2.27, 95% Confidence Interval 1.15-4.45, *P*=0.02).

**Conclusion:** Insulin dependence is associated with worse cancer-specific survival in PNET patients following surgical resection compared to PNET patients with NIDDM or without DM.

## Introduction

Pancreatic neuroendocrine tumors (PNET) are rare tumors arising from the islet cells of the pancreas [1]. Although the incidence remains low at roughly 2-3 per 100,000 annually in the United States (US) [1–3], the overall incidence has been increasing over the last two decades as small, nonfunctional PNETs are more frequently diagnosed due to the widespread use of cross-sectional imaging [4].

While their clinical courses are remarkably dissimilar, PNETs and pancreatic adenocarcinoma (PDAC) share a likeness in that they both show a strong concurrence with diabetes mellitus (DM) [5,6]. As many as 25-55% of PNET patients have coinciding DM at the time of PNET diagnosis compared to 7-9% of the general US population [7,8]. Some studies suggest that pre-existing DM is a risk factor for the development of PNETs and that a concurrent diagnosis of DM is associated with poor prognosis in PNET patients, without distinction between insulin dependent (IDDM) and non-insulin dependent DM (NIDDM) [6,9,10]. Evidence from the multicenter Pancreatic Retrospective Italian Metformin-NET (PRIME-NET) study analyzing patients with advanced PNETs treated both medically and surgically showed that metformin use was significantly associated with improved progression-free survival outcomes compared to non-diabetics as well as diabetics not taking metformin, including those using insulin and other hypoglycemic agents [11]. Additionally, one recent publication suggested that poor preoperative glycemic control, as indicated by elevated preoperative hemoglobin A1c (HbA1c), is strongly associated with elevated PNET growth rates [12]. Although the mechanism is not entirely understood, all of this evidence suggests that metabolic disease characteristics of DM are likely to influence PNET progression and, ultimately, PNET patient outcomes.

Additionally, exogenous insulin use has been associated with poor outcomes in several malignancies, including breast, colon, and prostate cancers, as well as in PDAC [13–18], the impact of insulin dependence in PNET patients remains unexamined. The primary aim of this study was to utilize a large, multi-institutional retrospective database to evaluate insulin dependence as a predictor of cancer-specific survival among PNET patients.

## Methods

### Data Collection and Study Design

The US Neuroendocrine Tumor Study Group (US-NETSG) is a collaboration of eight academic medical centers: Vanderbilt University, Emory University, Stanford University, The Ohio State University, Stanford University, Virginia Mason University, and Washington University in St. Louis, University of Michigan, and University of Wisconsin. Adult patients who underwent resection of gastro-entero-pancreatic neuroendocrine tumors between 2000 and 2016 were retrospectively identified at each institution. The institutional review board approved the data collection of each participating institution.

Over 750 parameters were collected in the database for each patient, including but not limited to demographics, general health information and comorbidities, preoperative imaging and laboratory data, intraoperative data, pathology data, postoperative complications, time to recurrence/progression/death, and cause of death. The American Joint Commission on Cancer Staging Manual, seventh edition [19], was used to define the TNM classification. Survival data and causes of death were determined through chart review. The de-identified data from each institution were shared among the collaborating institutions for analysis.

All patients who underwent PNET resection for a reported cause of death were included in the analysis. A documented preoperative diagnosis defined diabetic status (non-DM vs. DM). DM patients were further classified as either insulin-dependent (IDDM) if they used exogenous insulin preoperatively or non-insulin dependent (NIDDM) if they did not. Cancer-specific survival (CSS) was calculated as the time from index operation to cancer-specific death. Demographic, clinical, and tumor pathology parameters, as well as CSS, were compared across all three study arms (IDDM vs. NIDDM vs. Non-DM) and among those who did and did not report preoperative insulin therapy use (IDDM vs. NIDDM + Non-DM).

### Statistical Analysis

Demographic, preoperative, intraoperative, pathologic, and survival data were compared according to insulin dependence and diabetic status. Categorical variables were presented as frequencies and percentages and compared using chi-square or Fisher’s exact test, as appropriate. Continuous variables were reported as median values with interquartile ranges (IQR) unless otherwise specified and compared using the Kruskal-Wallis test. DSS was calculated using the Kaplan-Meier method and compared using the log-rank test. Clinical and pathological data were analyzed using multivariate Cox regression methods. Significance was set at a *p-value* < 0.05. All statistical analyses were performed using 1.1.383 R statistics software (R Core Team Vienna, Austria).

## Results

Of the 1,247 patients who underwent primary PNET resection during the study period (2000-2016) at any of the US-NETSG participating institutions, 1,122 (90%) had both preoperative presence or absence of DM and cause of death recorded and were included in our analysis. Within the study cohort, 870 (77%) were non-DM, 168 (15%) had NIDDM, and 84 (8%) had IDDM at the time of PNET resection. The groups were similar in race and operative intent (curative vs. non-curative), primary tumor size, TNM clinical stage, and tumor grade. The demographics, clinical, and tumor pathology parameters of the three sub-groups (non-DM, NIDDM, and IDDM) are listed in Table 1. The groups differed in sex, BMI, age, ASA class, preoperative HbA1c and serum glucose, and intrapancreatic location, as well as rates of preoperative hypertension, previous coronary events, and dyspnea symptoms. Among these factors, ASA class was the only factor associated with shortened CSS and a potential confounder. In comparison, according to insulin-dependence (IDDM vs. NIDDM + No-DM), IDDM patients had significantly higher levels of HbA1c%, serum glucose, and rates of hypertension, previous coronary events, and tumors within the body and tail of the pancreas. A complete list of comorbidity rates and univariate survival analysis are summarized in Supplementary Table 1 and Supplementary Table 2, respectively.

**Table 1.**
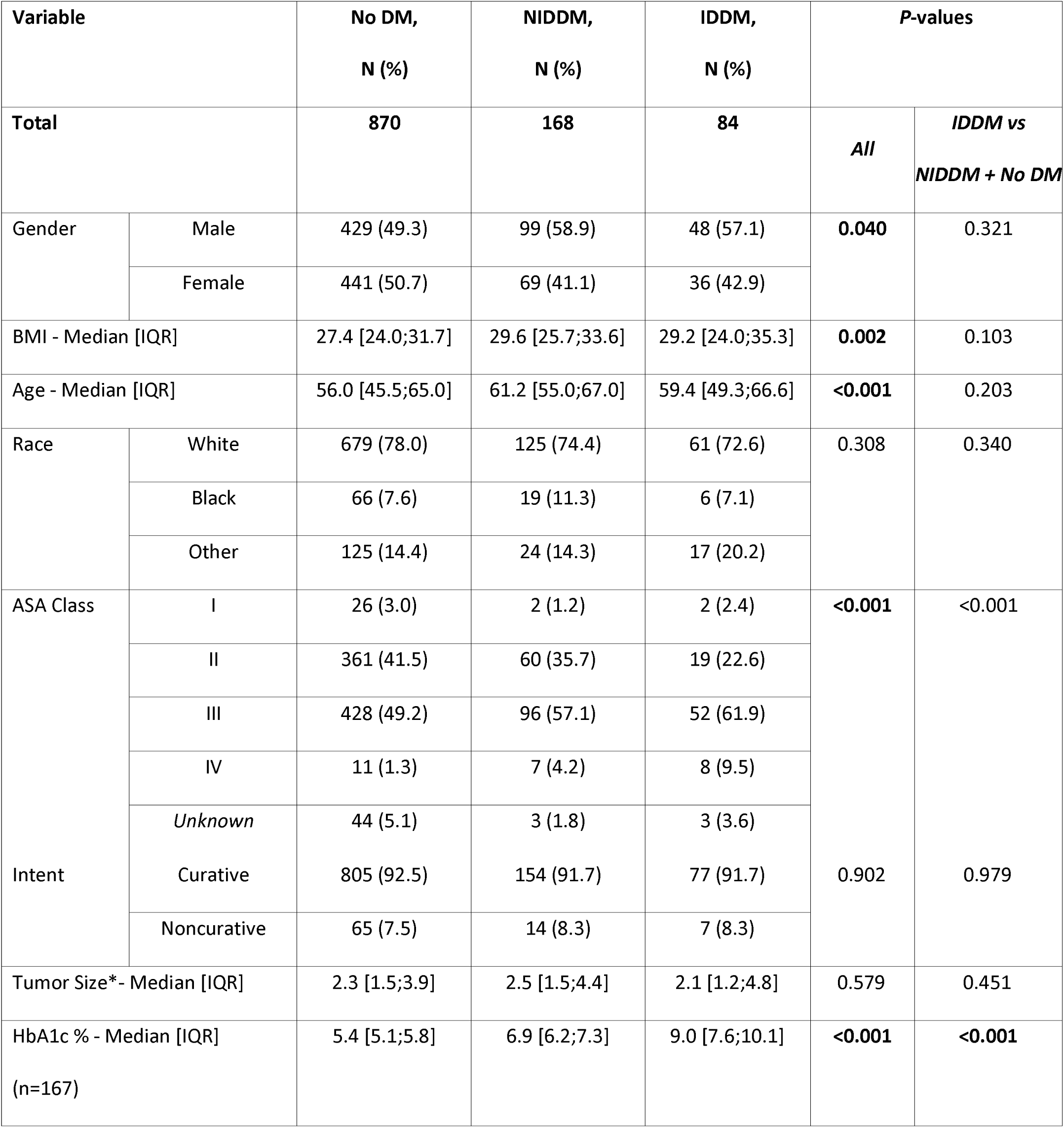
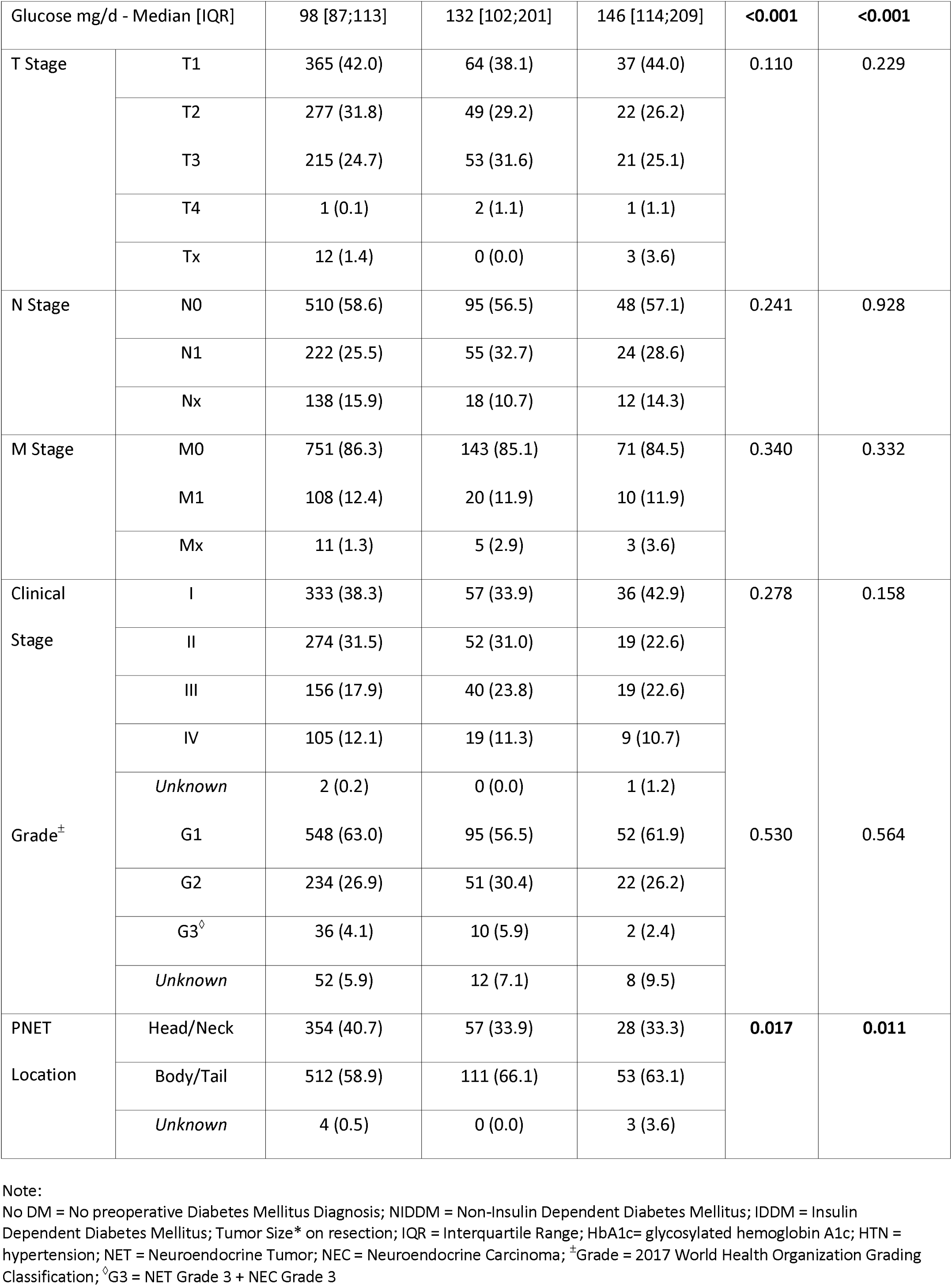
Demographics, clinical, and tumor pathology parameters.

Patients with IDDM had significantly decreased 5-year CSS compared to non-DM and NIDDM patients (CSS: IDDM 85%, NIDDM 94%, non-DM 93%, *P* <0.01; NIDDM + non-DM 93%, *P*<0.01) (shown in Fig. 1. a-b). The median CSS was not met for any group. In CSS multivariable analysis, after accounting for ASA class, factors associated with worse CSS included ASA class IV and insulin dependence. Insulin dependence was independently associated with worse CSS (Hazard Ratio 2.27, 95% Confidence Interval 1.15 – 4.45, *P*=0.02) than patients without DM (Table 2).

**Figure 1.**
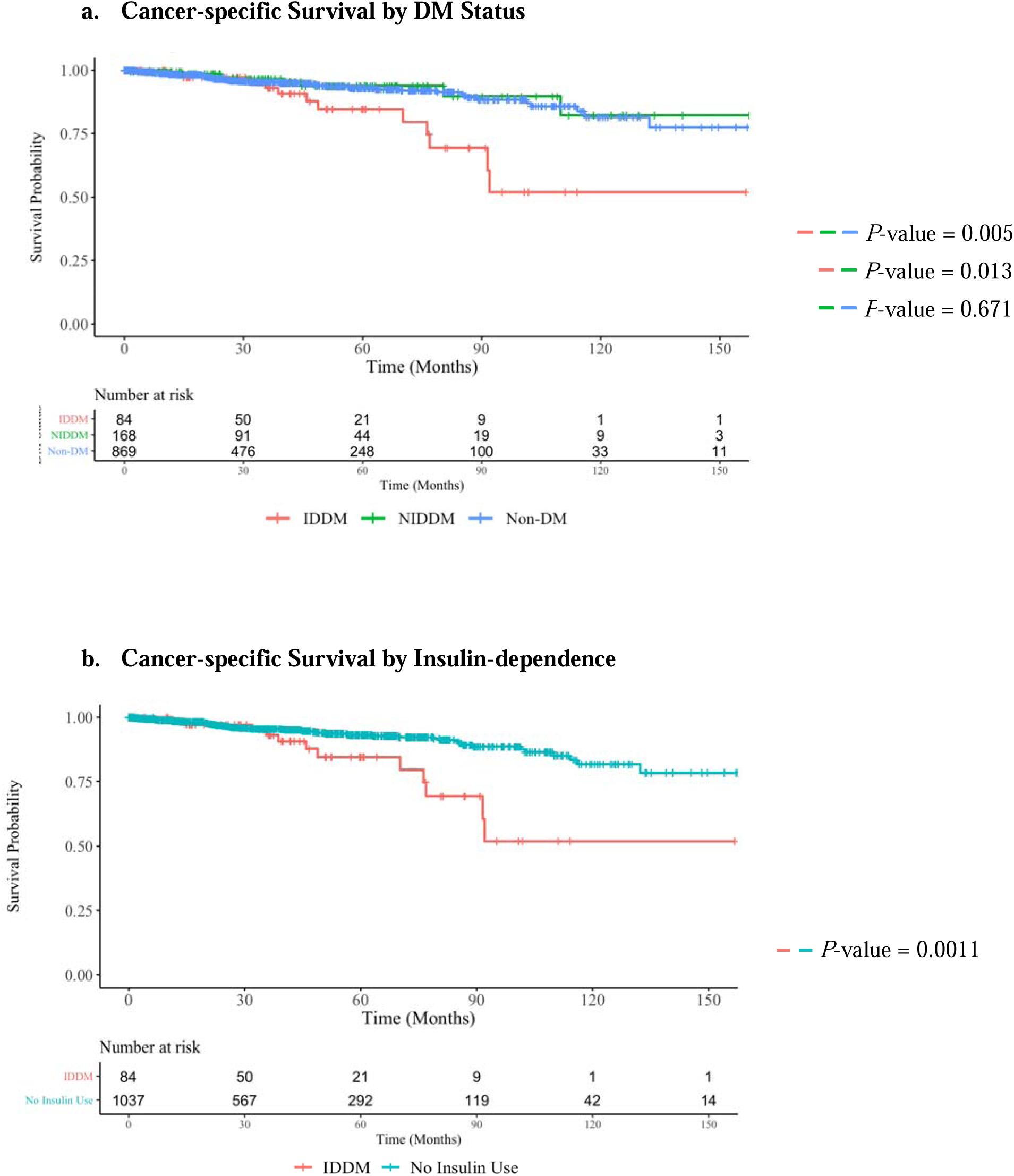
Disease-specific Kaplan-Meier Survival analysis

**Table 2.**
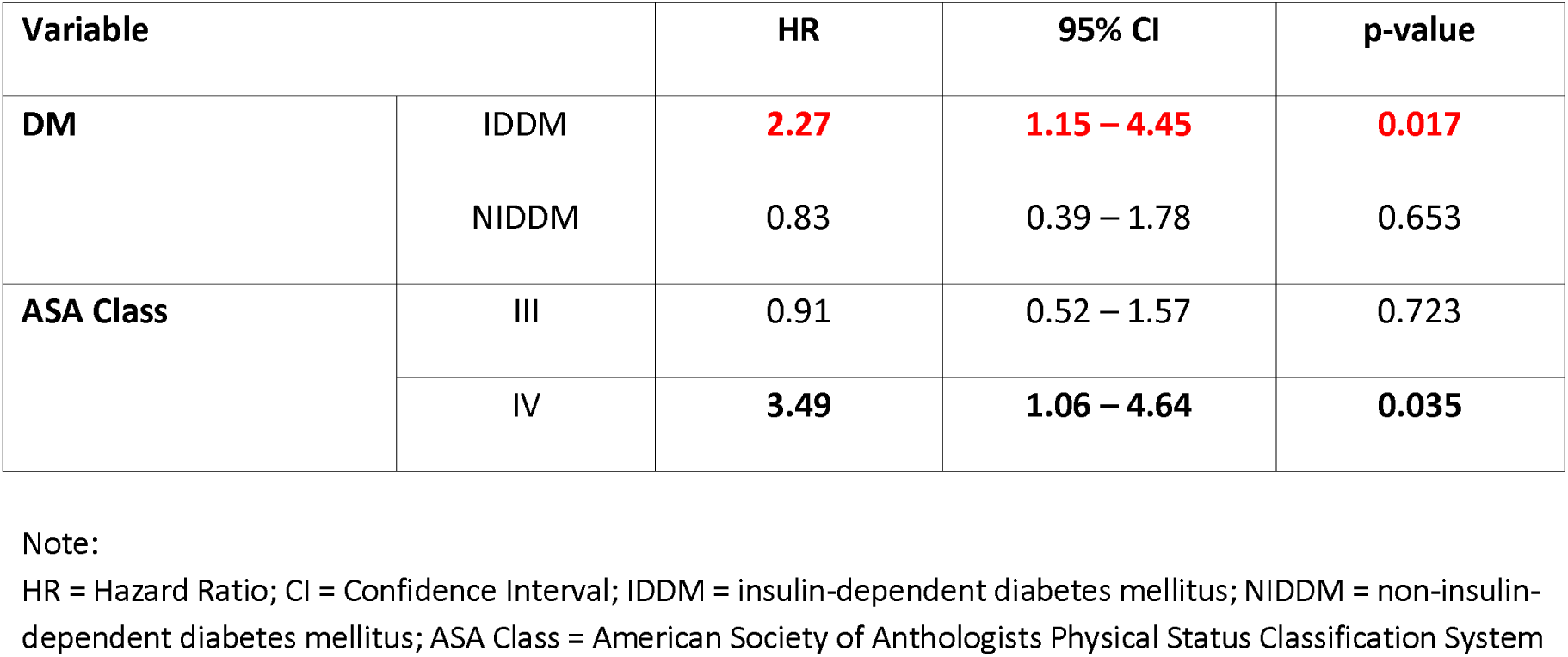
Cancer-specific Survival Cox Multivariate Analysis.

## Discussion

The current study identified a significant correlation linking insulin dependence to worse CSS following surgical PNET resection. On multivariable analysis, insulin dependence was further observed as an independent risk factor for CSS compared to NIDDM and non-DM PNET patients. The higher rate of cancer-specific mortality, despite more favorable tumor stage and grade, in such a generally indolent disease raises concern and the opportunity for improved prognostication and clinical management of PNET patients with IDDM.

Evidence demonstrating insulin-induced carcinogenesis and mitogenicity, as well as increased mortality, has been demonstrated in malignancies, including breast, pancreatic, colorectal, and prostate cancers [13–18]. We believe the associations identified in this study, however, to be particularly compelling in light of recent literature describing the mechanisms of PNET progression as well as the high prevalence of IDDM among PNET patients, which steadily increased through the duration of this study (shown in Fig. 2) [20–24]. In PNETs, insulin/IGF-1 signaling cascades (PI3K-Akt-mTOR, Ras/Raf/MEK/ERK) are well-established drivers of malignant islet cell proliferation [25–27]. Since the RADIANT-1 and 3 clinical trials demonstrated the efficacy of the mTOR inhibitor Everolimus in hindering the progression of advanced PNETs in 2011 [28,29], these anabolic pathways have become standard targets of medical treatments and combinations [30–35]. In 2016, Pusceddu and Vernieri et al. identified improved progression-free outcomes among PNET patients with concurrent type 2 DM (T2DM) taking metformin, a widely used oral hypoglycemic agent with antineoplastic properties attributed to indirect mTOR inhibition, compared to non-DM and DM patients with alternative DM management (including insulin therapy) [11,36–38].

**Figure 2.**
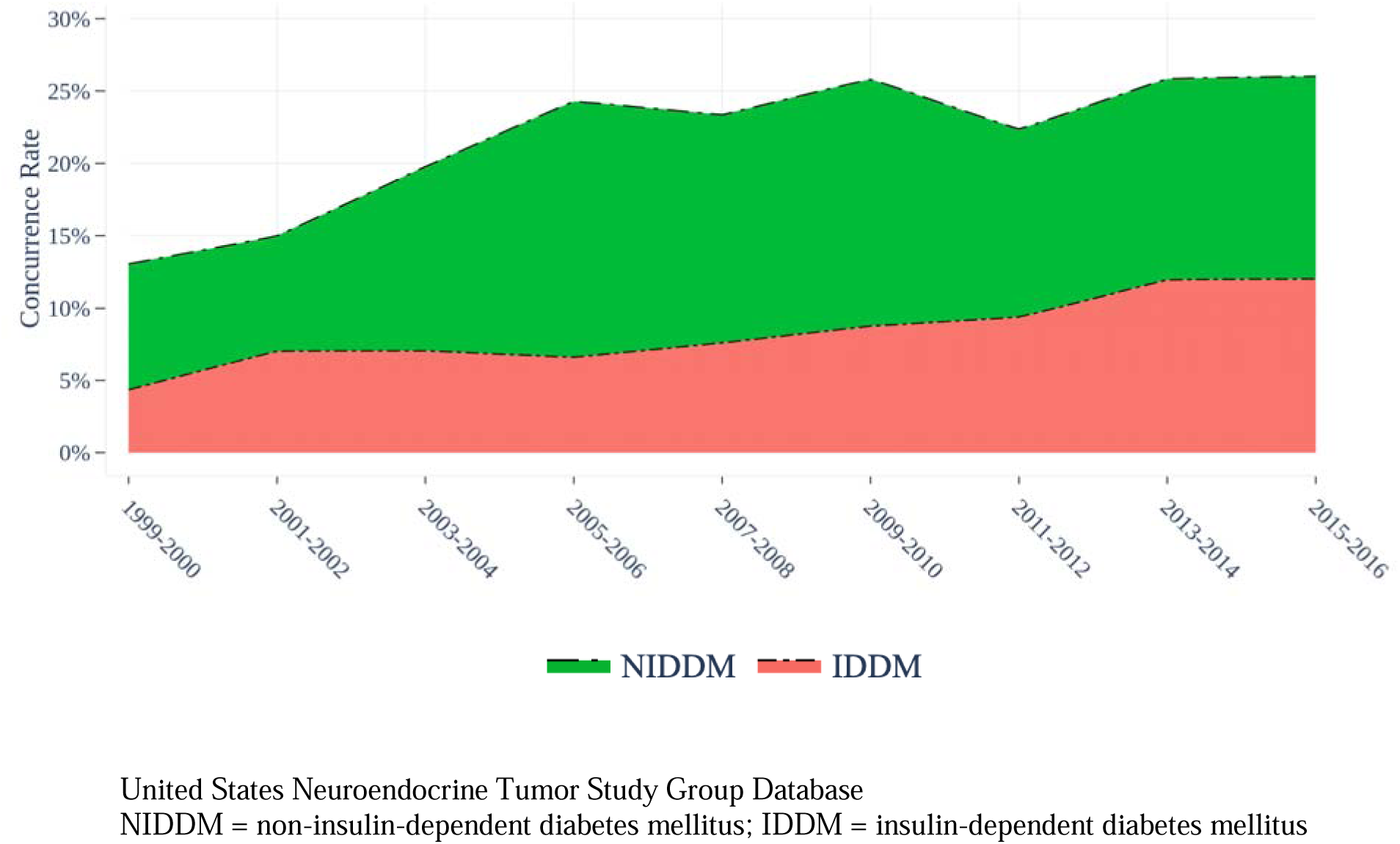
Concurrence of Pancreatic Neuroendocrine Tumors and Diabetes Mellitus

More recently, Vernieri and Pusceddu et al. determined that the poor outcomes among PNET patients with DM were attributable to metabolic disease after observing hypertriglyceridemia and hypercholesterolemia to be independently associated with shortened progression-free survival after accounting for metformin use. In contrast, hyperglycemia measured by elevated glycosylated hemoglobin A1c % (HbA1c%) was not [39].

In a previous USNET-SG database study, Baechle et al. identified elevated HbA1c% as significantly associated with an elevated tumor-specific growth rate [12]. However, HbA1c has been consistently not found to be independently associated with worse PNET prognosis in these prior studies [11,12,40], nor was it in this study. Considering this collage of evidence, our findings suggest that either the underlying metabolic derangements characteristic of IDDM (including hypertriglyceridemia, hypercholesterolemia, and hyperinsulinemia), the use of exogenous insulin therapy, or both may conspire favorable PNET outcomes.

It is also possible that the relationship observed between IDDM and poor PNET prognosis may be attributable to endocrine disruptions secondary to the tumor mass effect or paraneoplastic inflammation. Although the tumor sizes were similar, tumors of the diabetic groups were disproportionally located in the body and tail regions of the pancreas in comparison to the head, neck, and uncinate. Because β-cell concentration is highest in the body and tail of the pancreas, tumors of these regions may inordinately disrupt β-cell function and insulin secretion [41]. This relationship between PNET location and DM has not been previously characterized and is inverse to that of PDAC [42]; however, these local and systemic phenomena are unlikely to fully explain the difference in cancer-specific survival.

Of course, the retrospective nature of our study limits our ability to draw conclusions relating to causality. However, we also acknowledge that this study carries several other limitations. First, the US-NETSG database only classified DM patients as insulin-dependent, oral therapy, and diet-controlled DM, and therefore, does not delineate between type 1 and type 2 DM; however, due to the predominance of T2DM in the United States and its strong concurrence with PNETs, we believe this to have little impact on our findings. This report also does not specify insulin type (long-acting vs. short-acting), oral medication (metformin vs. sulfonylurea), or combined therapy regimens. Recent studies attributed decreased tumor progression and superior outcomes to metformin use over other T2DM oral medications in PNET patients, as well as those with other malignancies (breast, pancreatic, colorectal, prostate) [43–47]. Furthermore, long-acting, high-affinity insulin analogs, particularly glargine, have been associated with higher incidence, more rapid tumor progression, and inferior outcomes among several cancers (breast, pancreatic, colorectal, prostate) compared to the use of Neutral Protamine Hagedorn (NPH) insulin and other insulin analogs in the treatment of T2DM [14–18]. However, unfortunately, due to the limitations of the US-NETSG database, it is not possible to conduct a comparative sub-analysis of IDDM and NIDDM being treated with one insulin type or oral DM agent over another, and we acknowledge this as an area of interest for future studies.

Second, the DM status of this cohort was determined by chart review of the preoperative period and, therefore, cannot account for patients who may have recently altered treatment plans preoperatively or those whose DM status or management may have changed following their resection [particularly in the case of patients with total pancreatectomies, n=21 (2%)]. Third, because the retrospective database includes only those patients who underwent PNET resection, we failed to capture those patients being treated non-operatively, including those with small tumors being observed and those with unresectable malignancy. Next, though the multi-institutional collaboration behind this study allowed us to assemble a robust database characterizing this rare tumor, the consortium was limited to large academic referral centers, which may lead to selection bias and limit the generalizability of our conclusions in other populations. Lastly, the US-NETSG database is somewhat limited in terms of data related to tumor biology, including data on tumor grade, Ki-67, mitotic index, differentiation, serum insulin levels, and HbA1c%, and did not report lipid panel lab values (triglycerides, cholesterol, lipoproteins, cholesterol). Because such lab tests, specifically lipid panels, serum insulin, and HbA1c%, are not routine lab tests for all patients, they are unlikely to be measured pre-operatively without clinical indication or suspicion of pre-existing DM diagnosis or insulinoma. Therefore, we could not fully examine these factors related to diabetic status and PNET patient outcomes in our patient cohort. Similarly, the scarce reporting of tumor Ki-67 and mitotic rate limited our study from further delineating the possible correlations between DM therapy and tumor biology. Similarly, the limited reporting of HbA1c% and lack of lipid lab values hindered comparative analysis between hypertriglyceridemia, hypercholesterolemia, and hyperglycemia as to how these factors may collectively or individually impact PNET progression and patient prognosis, as postulated by previous studies [48]. Furthermore, elevated HbA1c was not associated with shortened CSS in the sub-analysis. However, because of our limited capacity to control for chronic hyperglycemia as a potential confounder, we did not attempt overall survival analysis comparative to DM status because HbA1c is widely accepted as a risk factor for diabetes-related mortality. Therefore, our analysis focused on cancer-specific death, where HbA1c% is not a direct confounder in PNET oncologic outcomes [11,12,39].

Despite these limitations, our study provides further insight into the interplay between PNETs and DM and how the insulin-dependent subset of PNET-DM patients has a uniquely inferior cancer prognosis. Additionally, the concurrence of DM among PNETs rose twofold (13% to 26%) and that of IDDM increased almost threefold (4.5% to 12%) throughout this study (1999-2016), suggesting a growing clinical challenge and need for the investigation of optimal co-management of DM and IDDM (shown in Fig. 2).

## Conclusion

In conclusion, although DM has previously been shown to be a risk factor for worse outcomes in PNET patients, the disparity in survival outcomes associated with insulin dependence among PNET patients with DM has not been previously described. When delineating DM by insulin dependence, IDDM alone appears to be associated with a worse cancer prognosis following surgical resection. Should a causal relationship exist between insulin therapy and PNET cancer-specific death, preferencing methods of peripheral insulin sensitization over insulin therapy in certain patients with PNET may have the potential to alter oncological outcomes in PNET differentially. However, in the case of a non-casual but co-linear relationship between DM and PNET severity, IDDM may indicate the clinical course, supplementing standard stage and grade prognostication.

## Acknowledgement

None

## Statement of Ethics

The protocol for this study was reviewed and approved by Vanderbilt University Medical Center Institutional Review Board, approval number 170730.

## Author Contributions

MBM, JBJ, PMS, and DA wrote the original draft. JBJ curated the data and performed a formal analysis. MBM also presented the work. MBM, PMS, and DA reviewed and edited the manuscript. MD, GP, FGR, CSC, ERW, RCF, SKM, and KI conceptualized the topic, helped develop the study design, provided validation, conducted the investigation, and helped with resources. KI provided project administration and supervision.

## Data Availability Statement

Due to institutional policy, the data supporting this study’s findings are not publicly available but can be obtained from the corresponding author upon reasonable request.

