## Supplementary Table 1 for "Insulin-dependence as a Predictor of Shortened Cancer-specific Survival in Pancreatic Neuroendocrine Tumors: A Multi-Institutional Study from the United States Neuroendocrine Study Group"

| **Supplementary Table 1.** Pre-operative IDDM-related Comorbidities by Insulin-dependence | | | | | | |
| --- | --- | --- | --- | --- | --- | --- |
| **Variable** | | **No DM,**  **N (%)** | **NIDDM**  **N (%)** | **IDDM**  **N (%)** | ***P*-value** | |
| **Total** | | **870** | **168** | **84** | ***All*** | ***IDDM vs***  ***NIDDM + No DM*** |
| Hypertension | Yes | 317 (36.4) | 110 (65.5) | 53 (63.1) | **<0.001** | **<0.001** |
|  | No | 534 (61.4) | 57 (33.9) | 31 (36.9) |  |  |
|  | *Unknown* | 19 (2.2) | 1 (0.6) | 0 (0.0) |  |  |
| Previous Coronary Event | Yes | 48 (5.5) | 13 (7.7) | 14 (16.7) | **0.001** | **0.002** |
|  | No | 801 (92.1) | 154 (91.7) | 70 (83.3) |  |  |
|  | *Unknown* | 21 (2.4) | 1 (0.6) | 0 (0.0) |  |  |
| Chronic Heart Failure | Yes | 7 (0.8) | 1 (0.6) | 2 (2.4) | 0.245 | 0.126 |
|  | No | 845 (97.1) | 166 (98.8) | 82 (97.6) |  |  |
|  | *Unknown* | 18 (2.1) | 1 (0.6) | 0 (0.0) |  |  |
| Dyspnea | Yes | 16 (1.8) | 3 (1.8) | 4 (4.8) | **0.176** | 0.087 |
|  | No | 835 (96.0) | 164 (97.6) | 80 (95.2) |  |  |
|  | *Unknown* | 19 (2.2) | 1 (0.6) | 0 (0.0) |  |  |
| Acute Renal Failure | Yes | 0 (0.0) | 1 (0.6) | 0 (0.0) | 0.121 | 0.436 |
|  | No | 852 (97.9) | 166 (98.8) | 84 (100) |  |  |
|  | *Unknown* | 18 (2.1) | 1 (0.6) | 0 (0.0) |  |  |
| Chronic Renal Failure | Yes | 7 (0.8) | 1 (0.6) | 0 (0.0) | 0.053 | 0.126 |
|  | No | 845 (97.1) | 166 (98.8) | 83 (98.8) |  |  |
|  | *Unknown* | 18 (2.1) | 1 (0.6) | 1 (1.2) |  |  |
| Ascites | Yes | 1 (0.1) | 1 (0.6) | 1 (1.2) | 0.091 | 0.133 |
|  | No | 851 (97.8) | 166 (98.8) | 83 (98.8) |  |  |
|  | *Unknown* | 18 (2.1) | 1 (0.6) | 0 (0.0) |  |  |
| Other Malignancy | Yes | 37 (4.3) | 15 (8.9) | 3 (3.6) | 0.072 | 0.595 |
|  | No | 816 (93.8) | 152 (90.5) | 81 (96.4) |  |  |
|  | *Unknown* | 17 (2.0) | 1 (0.6) | 0 (0.0) |  |  |
| Disseminated Cancer | Yes | 57 (6.6) | 15 (8.9) | 10 (11.9) | 0.120 | 0.145 |
|  | No | 795 (91.4) | 152 (90.5) | 74 (88.1) |  |  |
|  | *Unknown* | 18 (2.1) | 1 (0.6) | 0 (0.0) |  |  |
| Identified Genetic Mutation | Yes | 89 (10.2) | 12 (7.1) | 11 (13.1) | 0.063 | 0.382 |
|  | No | 764 (87.8) | 155 (92.3) | 73 (86.9) |  |  |
|  | *Unknown* | 17 (2.0) | 1 (0.6) | 0 (0.0) |  |  |
| Pancreatitis | Yes | 43 (4.9) | 13 (7.7) | 9 (10.7) | 0.063 | 0.076 |
|  | No | 808 (92.9) | 154 (91.7) | 75 (89.3) |  |  |
|  | *Unknown* | 19 (2.2) | 1 (0.6) | 0 (0.0) |  |  |
| Anemia | Yes | 63 (7.2) | 16 (9.5) | 8 (9.52%) | 0.249 | 0.449 |
|  | No | 787 (90.5) | 151 (89.9) | 76 (90.5%) |  |  |
|  | *Unknown* | 20 (2.3) | 1 (0.6) | 0 (0.00%) |  |  |
