## Supplementary Table 2 for "Insulin-dependence as a Predictor of Shortened Cancer-specific Survival in Pancreatic Neuroendocrine Tumors: A Multi-Institutional Study from the United States Neuroendocrine Study Group"

**Supplementary Table 2.** Cancer-specific Survival Cox Univariate Analysis

| **Variable** | | | **HR** | **95% CI** | **p-value** |
| --- | --- | --- | --- | --- | --- |
| **DM** | **IDDM** | | **2.65** | **1.41 – 4.99** | **0.003** |
|  | NIDDM | | 0.86 | 0.41 – 1.81 | 0.685 |
| **Male** | | | 0.92 | 0.57 – 1.47 | 0.713 |
| **Age** | | | 1.01 | 0.99 – 1.03 | 0.475 |
| **BMI** | | | 0.95 | 0.90 – 0.99 | **0.028** |
| **ASA Class** | | III | 0.93 | 0.54 – 1.60 | 0.793 |
|  |  | **IV** | **4.53** | **1.84 – 11.1** | **<0.001** |
| **HbA1c (n=167)** | | | 1.41 | 0.90 – 2.20 | 0.134 |
| **Serum Glucose** | | | 1.00 | 0.99 – 1.01 | 0.284 |
| **PNET Location: Head/Neck** | | | 1.14 | 0.71 – 1.85 | 0.586 |
| **Hypertension** | | | 1.03 | 0.63 – 1.70 | 0.904 |
| **Previous Coronary Event** | | | 0.56 | 0.13 – 2.31 | 0.427 |
| **Dyspnea** | | | 1.07 | 0.15 – 7.71 | 0.950 |

HR = Hazard Ratio; CI = Confidence Interval; DM = diabetes mellitus; IDDM = Insulin dependent diabetes mellitus, NIDDM = non-insulin dependent diabetes mellitus; HbA1c = glycosylated hemoglobin A1c
